# Assessing Hot Water Reconstitution Practices and Labeling of Powdered Infant Formula to Enhance Microbial Safety Involving Cronobacter spp

**DOI:** 10.1101/2025.03.21.25324421

**Authors:** Maria Amalia Beary, Sarah E. Daly, Jakob Baker, Abigail B. Snyder

## Abstract

*Cronobacter* spp. contamination in powdered infant formula (PIF) can cause infections in high-risk infants. Public health guidelines for caregivers of high-risk infants advise reconstitution using water heated to at least 70°C (158°F) for microbial inactivation. This study evaluated changes to water temperature under different heating and cooling scenarios during formula preparation, aiming to identify which conditions best ensure a minimum treatment temperature of 158°F (70°C). Factors such as vessel type, lid usage, vessel removal from the heat source, and water volume were tested for their effects on heat retention. The popular “hot shot” method which uses a small volume of hot water followed by a larger volume of room temperature water was also evaluated. In many scenarios, water temperatures fell below 158°F (70°C) during the preparation steps prior to PIF reconstitution. The water temperature prior to transfer to the bottle, bottle material, capacity, and volume significantly impacted temperature (*p*< 0.001). The temperature of formula immediately following shaking was as high as 179.5±1.6°F (81.9±0.9°C) and as low as 138.0±1.3°F (58.9±0.7°C), depending on the preparation conditions. Bottle material and capacity, water volume, and initial water temperature significantly impacted the temperature of PIF reconstitution treatments. Small volumes (2 fl. oz) of water in small glass bottles cooled the quickest. The hotshot method yielded reconstitution temperatures below 158°F (70°C). Measuring the temperature of hot water in the baby bottle and adding PIF when it cooled to 165°F (73.8°C) resulted in formula temperatures at or above 158°F (70°C) in almost all cases. Labels from PIF products available in the U.S. were reviewed and lacked detailed information about how caregivers of high-risk infants should use hot water to reconstitute PIF. The findings of this study can help shape guidelines that improve PIF reconstitution practices.

## INTRODUCTION

*Cronobacter* spp. infections in high-risk infants, typically defined as infants under 2 months of age, born prematurely, or otherwise immunocompromised, can result in serious adverse health consequences including septicemia, meningitis, and death (WHO/FAO 2006, CDC, 2024). Although most infections are caused by *C. sakazakii,* three additional species (*C. malonaticus*, *C. turicensis,* and *C. dublinensis*) have been linked to clinical cases (Li et al, 2023). Powdered infant formula (PIF) is one vector through which infants can be infected with *Cronobacter* spp. PIF contamination can occur during manufacturing (CDC, 2002). *Cronobacter* spp. can tolerate osmotic stress and elevated temperatures which may contribute to their survival in dry food processing environments (Healy et al., 2010). *Cronobacter* spp. are also found in household settings and cross-contamination may occur during consumer handling (Samadpour et al., 2024).

Because of the risk of *Cronobacter* spp. contamination in PIF, public health agencies recommend additional precautionary measures for high-risk infants. One key recommendation is the use of hot water to reconstitute PIF, which is based on validation studies that show that hot water at 70°C (158°F) or above will instantaneously inactivate > 5 log CFU/ml *C. sakazakii* (Edelson-Mammel and Buchanan, 2004). For high risk infants, U.S. public health guidance generally suggests bringing water to a boil, then letting it cool in the heating vessel for no more than 5 min before addition into a clean bottle, then mixing in PIF (CDC, 2024; FDA, 2024). This guidance omits temperature measurements during formula preparation, which may increase convenience for the caregiver. However, these time-based instructions can also translate to inconsistency in water heating practices among caregivers.

Caregivers often rely on product labels for formula preparation instructions, though these labels may not provide guidance on PIF reconstitution with hot water (Malek et al., 2020). There are a variety of foreseeable formula preparation approaches that caregivers could use which reasonably result in vastly different water temperatures at different stages of preparation. As one example, the “hot shot” method in which only 1 oz (30 mL) of water at 70°C (158°F) is initially added to PIF, may not be sufficient to inactivate *Cronobacter* spp. Additionally, the 70°C (158°F) water temperature is typically indicated in the water heating vessel (e.g., stainless steel pot, electric kettle) *prior* to transfer to the baby bottle. This fails to consider the inevitable heat loss during the water transfer process. Additionally, recommendations based on a holding time neglect how different heating vessel materials and environmental conditions impact heat loss during the 5 min cooling period. There has been little investigation into variations in temperature at the end of a 5 min cooling period in different heating vessels, and at the point of reconstitution. This study aimed to 1) quantify the differences in water temperature outcomes based on reasonably foreseeable preparation conditions, 2) identify preparation parameters that result in water temperatures below 70°C (158°F) during PIF reconstitution, and 3) evaluate temperature and safe-handling recommendations within the preparation instructions of commercial PIF products.

## MATERIALS AND METHODS

### Heating Vessels

Three vessel types were selected to boil (100°C; 212°F) water for preparation of PIF, including: (i) 1.5 L stainless steel pot (Winco® SSSP-2 #47, Lodi, NJ, USA), (ii) 1.7 L stainless steel electric kettle (Phonect Electric Kettle Temperature Control KE4065T-GS, Shenzhen Aukeyhi Technology Co., Ltd., China.), and a (iii) 1 L borosilicate glass Pyrex cup (Corning, New York, USA). Each heating vessel type had different capacities, designs, and other attributes (Table 1)

**Table 1.**
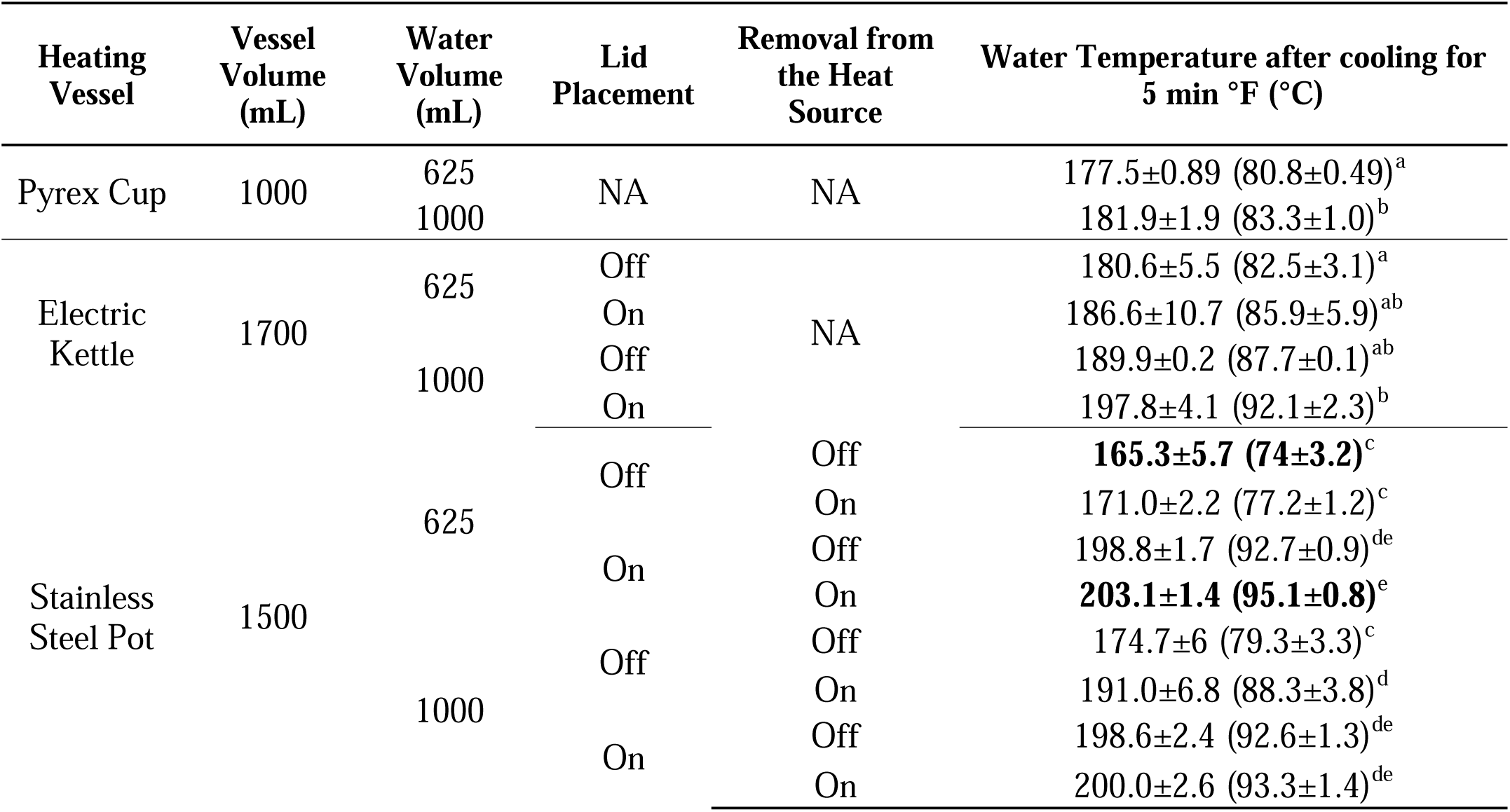
Water temperature within the heating vessels after cooling 5 min (Temperature Drop 1) in three types of heating vessels (Pyrex cup, Electric Kettle, Stainless Steel Pot). Letters indicate significantly different outcomes. Statistical tests were performed among results for each heating vessel. Results in bold text represent conditions that yielded the highest and lowest temperature among all conditions tested. Vessel volume (P<0.01), water volume (P<0.05), vessel removal from the heat source (P<0.01), and lid placement (P<0.001) significantly impacted water temperature.

### Water Volumes

Two volumes of water (1,000 mL and 625 mL) were introduced to each vessel (Table 1). Volumes were selected based on volume estimations for eight bottles a day (feeding infants ≤2 months, 125 mL/bottle as average) (La Leche League, 2022). When the heating vessels were filled with these two selected volumes (1,000 mL and 625 mL), the stainless steel pot was at 67% and 42% capacity, the electric kettle was at 59% and 37% capacity, and the Pyrex cup was at 100% and 62.5% capacity.

### Heat Sources

Different heating sources were used to heat the water in each vessel type (i.e, stainless-steel pot, electric kettle, and Pyrex cup). These heating sources were a gas-lit flame on the stove (for stainless steel pot), resistive heating via electricity flowing through the base heating element (for electric kettle), and a microwave oven (for Pyrex cup).

### Baby Bottle Materials

Baby bottles made of glass and plastic were used. Plastic bottles were Dr. Brown’s Natural Flow® BPA-free polypropylene plastic with Level 1 Slow Flow Nipple (St. Louis, Missouri) and glass, Dr. Brown’s Natural Flow® Anti-Colic Options+™ Narrow Glass (St. Louis, Missouri). Glass bottles were also from Lifefactory® borosilicate glass baby bottle with silicone sleeve, polypropylene cap, ring, and stopper, and a Stage 1 (0-3 months) silicone nipple (Sausalito, CA). Each baby bottle had different capacities (8 oz and 4 oz, each material), and dimensions (between 4.5cm and 5 cm of diameter, and between 4.5 and 6 inches of height) (Table 2).

**Table 2.** Water temperature after transfer to baby bottles (Temperature Drop 2). The highest and lowest temperature after cooling boiling water (Table 1, Temperature Drop 1) were selected as the initial water temperatures in this experiment. Letters indicate significantly different outcomes. Results in bold text represent conditions that yielded the highest and lowest water temperature among all conditions tested.

| Material Type | Bottle Volume oz (mL) | Water Volume oz (mL) | Water Temperature °F (°C) |  |
| --- | --- | --- | --- | --- |
|  |  |  | Initial Temp 165 (73.8) | Initial Temp 203 (95.1) |
| Glass | 4 (118.3) | 2 (59.1) | **149.3±0.9 (65.2±0.5)<sup>a</sup>** | 165.7±1.3 (74.3±0.7)<sup>c</sup> |
|  |  | 4 (118.3) | 154±2.4 (67.8±1.3)<sup>b</sup> | 176.1±5.7 (80.1±3.2)<sup>d</sup> |
|  | 8 (236.6) | 4 (118.3) | 149.3±0.7 (65.2±0.4)<sup>a</sup> | 173.3±3.3 (78.5±1.8)<sup>c</sup> |
|  |  | 8 (236.6) | 153.1±0.5 (67.3±0.3)<sup>b</sup> | 181.2±4.1 (82.9±2.3)<sup>d</sup> |
| Plastic | 4 (118.3) | 2 (59.1) | 151±0.4 (66.1±0.2)<sup>ab</sup> | 174.5±1.6 (79.2±0.9)<sup>d</sup> |
|  |  | 4 (118.3) | 156.5±1.2 (69.2±0.7)<sup>b</sup> | 181.9±3 (83.3±1.7)<sup>e</sup> |
|  | 8 (236.6) | 4 (118.3) | 154.0±1.8 (67.8±1)<sup>ab</sup> | 179.2±1.1 (81.8±0.6)<sup>d</sup> |
|  |  | 8 (236.6) | 157.8±0.6 (69.9±0.3)<sup>b</sup> | **188.2±2.5 (86.8±1.4)<sup>e</sup>** |

### Procedures for heating, cooling, and transferring water

Water was heated to 100°C (212°F) in each heating vessel, then the heating sources were turned off. Water temperatures were measured continuously for up to 15 min (Table 1, Figure 1, 2). The use of a closed lid during the cooling process was evaluated, except for the Pyrex cup which is not designed to have a lid. Additionally, cooling in the stainless steel pot was evaluated when it was left on or removed from the heating element after the heat was turned off. Boiled water (100°C; 212°F) was cooled in the vessel for 5 min and then transferred to glass and plastic baby bottles (4 or 8 oz; 118.3 or 236.6 mL) (Figure 1, Table 2). The 5 min cooling duration was obtained from the CDC guidance to “bring water to boil and let it cool for not longer than 5 minutes” (CDC, 2024). Water temperatures in the baby bottle after the transfer were continuously monitored for 1 min. All experiments were conducted in triplicate.

**Figure 1.**
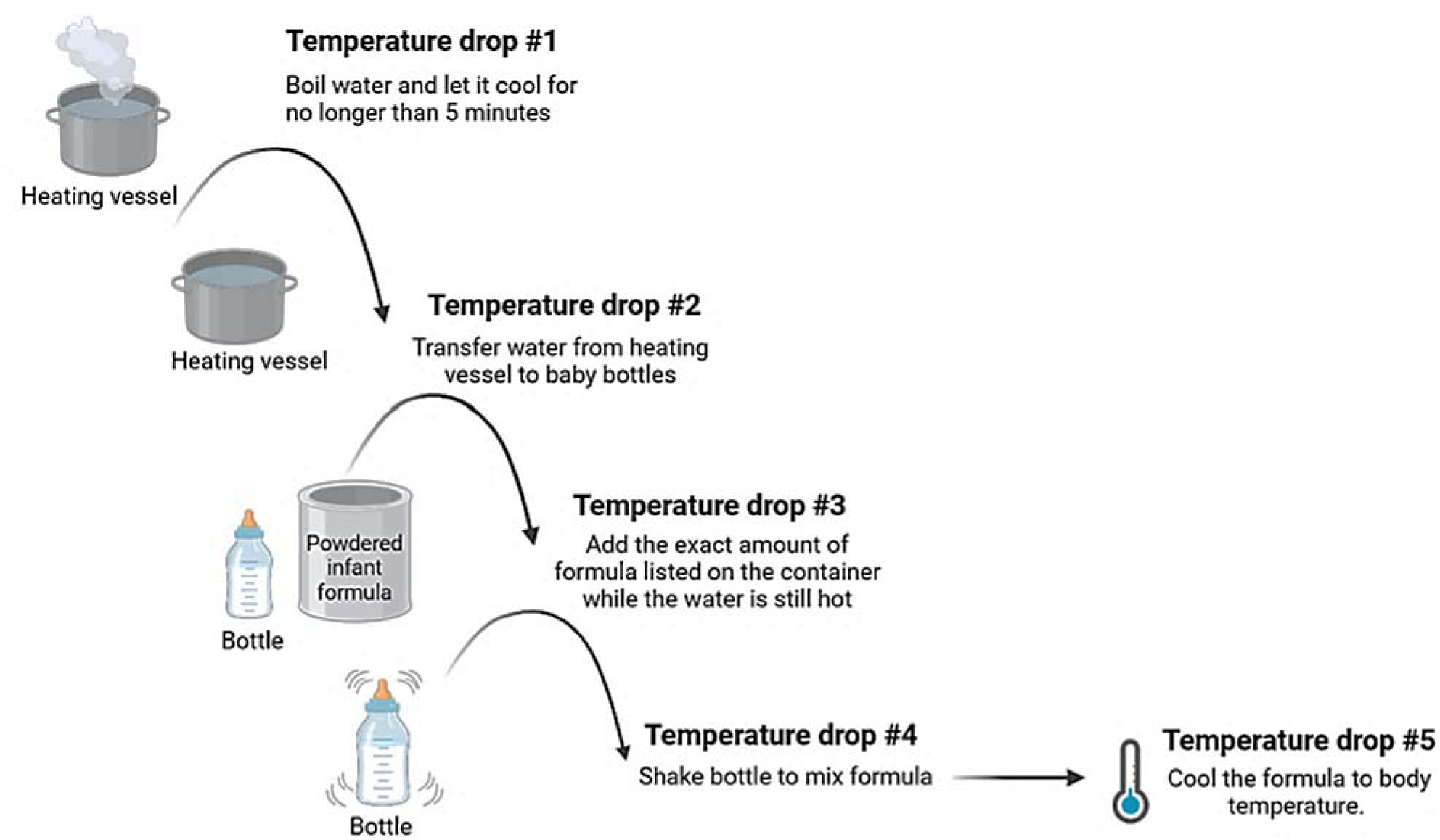
Sequential steps in the rehydration of PIF using hot water to enhance *Cronobacter* spp. control. The steps are derived from guidance documents published by the CDC (2024). Anticipated drops in water temperature are noted at each step.

In subsequent experiments, the highest and lowest temperatures achieved experimentally in the previous step of PIF reconstitution were used as initial temperatures. For example, following the trials evaluating water temperatures after 5 min of cooling, the resulting lowest and highest temperatures were 73.8°C (165°F) and 95.1°C (203°F), respectively. Therefore, these two temperature extremes were used as starting temperatures in the subsequent experiments evaluating temperature decreases following the transfer of the hot water to the baby bottles.

### Procedures for water temperature monitoring

Water temperatures were continuously recorded every 2 s with K-type thermocouples (Omega Engineering, Norwalk, CT) and an 8-channel handheld OM-HL-EH-TC data logger (OMEGA Engineering, Norwalk, CT, USA). Thermocouples were positioned at the (i) top-center, (ii) bottom-center, and (iii) center-edge of the vessels (Figure S1a). Preliminary results indicated that there was not a significant difference in water temperature among the three locations of the thermocouples (top-center, bottom-center, and center-edge), so temperatures measured at the top-center of vessels are reported in the results for all experiments (*p*>0.05). Within baby bottles, a thermocouple was positioned in the geometric center of the volume of water (Figure S1b). Location of thermocouple placement in the baby bottle also did not significantly impact temperature results.

### Powdered infant formula brand

One brand of PIF (Similac Total Comfort®, Abbott, OH) was used within the study. This product is a milk-based powdered infant formula with iron intended for infants 0-12 months of age. The ratio of formula/water (w/v) we used to reconstitute the formula was based on manufacturer’s instructions of 8.8 g per 2 fl. oz of water (Table S1).

### Formula Reconstitution

PIF (w/v: based on manufacturer’s guidelines) was introduced to the baby bottles after the heated water was transferred from the vessels to baby bottles (Figure 1, Table 3). We used water heated to 187.9°F (86.6°C) and 149.2°F (65.1°C), which were the highest and lowest temperatures obtained from the previous water heating experiment after cooling boiled water for 5 min in the heating vessel. Additional studies were conducted when the initial water temperatures in the baby bottles prior to PIF addition were: 73.8°C (165°F), 75°C (167°F), 76.6°C (170° F), 80°C (176°F), and 82°C (179.6°F).

**Table 3:** TOP Rows: Temperatures during PIF reconstitution with hot water and shaking (Temperature Drop 3 and 4). Initial water temperatures in this experiment were based on the highest and lowest water temperatures identified from previous steps (Table 1 and 2, Temperature Drop 1 and 2). Tests were conducted in bottle types and conditions that similarly presented the greatest and least heat retention. BOTTOM Rows: Temperatures during PIF reconstitution using the “hotshot” method in 4 oz bottles.

| Baby bottle material | Bottle volume oz (mL) | Total water volume oz (mL) | Volume heated to 70°C oz (g) | Temperature of added cold water °F (°C) | Initial temperature in vessel °F (°C) | Temperature when transfer to bottle °F (°C) | Temperature after Addition of PIF °F (°C) | Temperature after Addition of cold tap water °F (°C) | Shake 5s °F (°C) | Hold 5min °F (°C) | Condition |
| --- | --- | --- | --- | --- | --- | --- | --- | --- | --- | --- | --- |
| Plastic | 8 (236.6) | 8 (236.6) | NA | NA | 203 (95.1) | 187.9(86.6) | 182.1±1.7 (83.1±0.9) | NA | 179.5±1.6 (81.9±0.9) | 165.6±1.3 (74.2±0.7) | Best |
| Glass | 4 (118.3) | 2 (59.1) | NA | NA | 165 (73.8) | 149.2(65.1) | 143.4±2.0 (61.9±1.1) | NA | 138.0±1.3 (58.9±0.7) | 125.4±0.7 (51.9±0.4) | Worst |
| Plastic | 4 (118.3) | 4 (118.3) | 1 (17.6) | 64.0 (17.8) | 158 (70) | 150.4±0.8 (65.8±0.5) | 140.3±1.1 (60.2±0.6) | 93.3±4.0 (34.0±2.2) | NA | NA | Hotsh |
|  |  | 2 (59.1) | 1 (8.8) | 66.6 (19.2) | 158 (70) | 148.9±0.8 (64.9±0.5) | 139.5±2.5 (59.7±1.4) | 113.8±9.3 (45.4±5.1) | NA | NA | Hotsh |
| Glass | 4 (118.3) | 4 (118.3) | 1 (17.6) | 64.8 (18.2) | 158 (70) | 143.8±3.1 (62.1±1.7) | 128.1±6.7 (53.4±3.7) | 89.5±6.3 (32.0±3.5) | NA | NA | Hotsh |
|  |  | 2 (59.1) | 1 (8.8) | 58.8 (14.9) | 158 (70) | 145.9±2.4 (63.3±1.3) | 120.6±5.3 (49.2±2.9) | 112.6±1.6 (44.8±0.9) | NA | NA | Hotsh |

After the addition of PIF, baby bottles were immediately securely capped, held vertically, and shaken with repetitive motion along the vertical axis at a frequency of 2–3 shakes per s for a total duration of 5 s. This motion facilitated homogeneous mixing of the liquid and the reconstituted powder. Temperatures in the bottles were recorded during PIF addition, shaking, then waiting 5 min.

### The “hot shot” method

The “hot shot” method was evaluated as it is a consumer practice for rapid preparation of PIF (Grant, et al., 2023) (Table 3). Approximately 1 fl. oz (29.6 mL) of water was heated to 100°C (212°F), cooled to 70°C (158°F), then introduced to plastic and glass baby bottles (4 oz). Next, 8.8 g or 17.6 g of PIF was immediately added to the bottle, then swirled once. Cool water (17.5°C; 63.6°F) was added to the bottle until the desired final volume (2 or 4 fl. oz; 59.1 or 118.3 mL) was reached. Temperatures were continuously recorded throughout (i) water transfer to the bottles, (ii) PIF addition, and (iii) cool water introduction.

### Cooling methods for bottles containing prepared infant formula

Three methods for cooling reconstituted formula were evaluated: (i) passive cooling at room temperature, (ii) running cool water over the outside of the baby bottle, and (iii) utilizing an ice bath. Methods (ii) and (iii) are described in CDC (2024) recommendations to reduce cooling durations for prepared PIF. Temperatures were continuously recorded until the reconstituted formula reached body temperature (37°C), which follows CDC (2024) guidance.

### Statistical Analysis

All statistical tests were conducted with R-studio (version 4.2.2, R studio, Boston, MA, USA). Differences in temperatures based on heating conditions and baby bottle conditions were determined using Analysis of variance (ANOVA) and post-hoc Tukey’s test (multcomp package version) (Hothorn, 2008).

### Mixing instructions and recommendations on labels of powdered infant formula

PIF products (n=36) from nine brands manufactured in the U.S. and Europe were selected from commercially available products from retail stores (e.g., Target, CVS, Walgreens, Costco, BJs) within the U.S. Formulas were intended for infants ≤ 0 – 12 months old. Selected types of PIF included powders with standard composition, special formulas, and milk alternatives (i.e. hydrolyzed cow’s milk formula, amino acid-based formula, soy-based formula, etc.) (Verduci et al, 2019). Instructions on formula can labels (i.e., drawings, descriptions, and bolded text) were analyzed for their recommendations of water temperature for formula reconstitution. Namely, whether the label directions included guidelines for heating water ≥70°C (158°F). Other heating recommendations on labels were analyzed, including any instructions on microwave reheating and how to evaluate formula cooling. Additional information on storage conditions, resources for guidance on formula preparation, and sterilization practices were also assessed.

## RESULTS

### Heating and cooling conditions significantly influenced water temperature

Water temperature at the end of the 5 min cooling period varied by vessel type. The stainless steel pot had the largest range of temperatures among preparation conditions. After 5 min of passive cooling, the lowest average temperature (165.3±5.7°F, 74±3.2°C) occurred in the stainless steel pot when the water volume was 625 mL, there was no lid, and the pot had been removed from the heating surface. The highest average temperature (200± 2.6°F, 93.3 ± 1.4°C) occurred in the stainless steel pot when the water volume was 625 mL, the lid was on, and the pot remained on the heating surface. Water temperatures were significantly higher when stainless steel pots were covered with lids and cooled on the heating surface (*p*<0.01, Table 1, Figure 2). The average water temperatures in the electric kettles had a smaller range at 197.8±4.1°F (92.1±2.3°C) to 180.6± 5.5°F (82.5 ± 3.1°C) (Table 1). Electric kettles had higher average water temperatures even when the lid was open, most likely because the surface area of the kettle’s top opening was smaller than the stainless-steel pot’s, potentially restricting heat loss. A higher water volume significantly conserved water temperatures in the electric kettle with a difference of 17.2±4.8°F (9.6±2.7°C) between 1,000 mL and 625 mL (*p*<0.05) (Table 1). Water temperatures in the Pyrex cup were lower than water temperatures in the electric kettle. The 1,000 mL water volume in the Pyrex cup decreased to a temperature of 181.9±1.9°F (83.3±1.0°C), and the 625 mL volume was significantly lower (*p*<0.05) at 177.5±0.89°F (80.8±0.49°C). The electric kettle most effectively prevented temperature loss in water during the first 5 min of cooling, regardless of preparation conditions.

**Figure 2.**
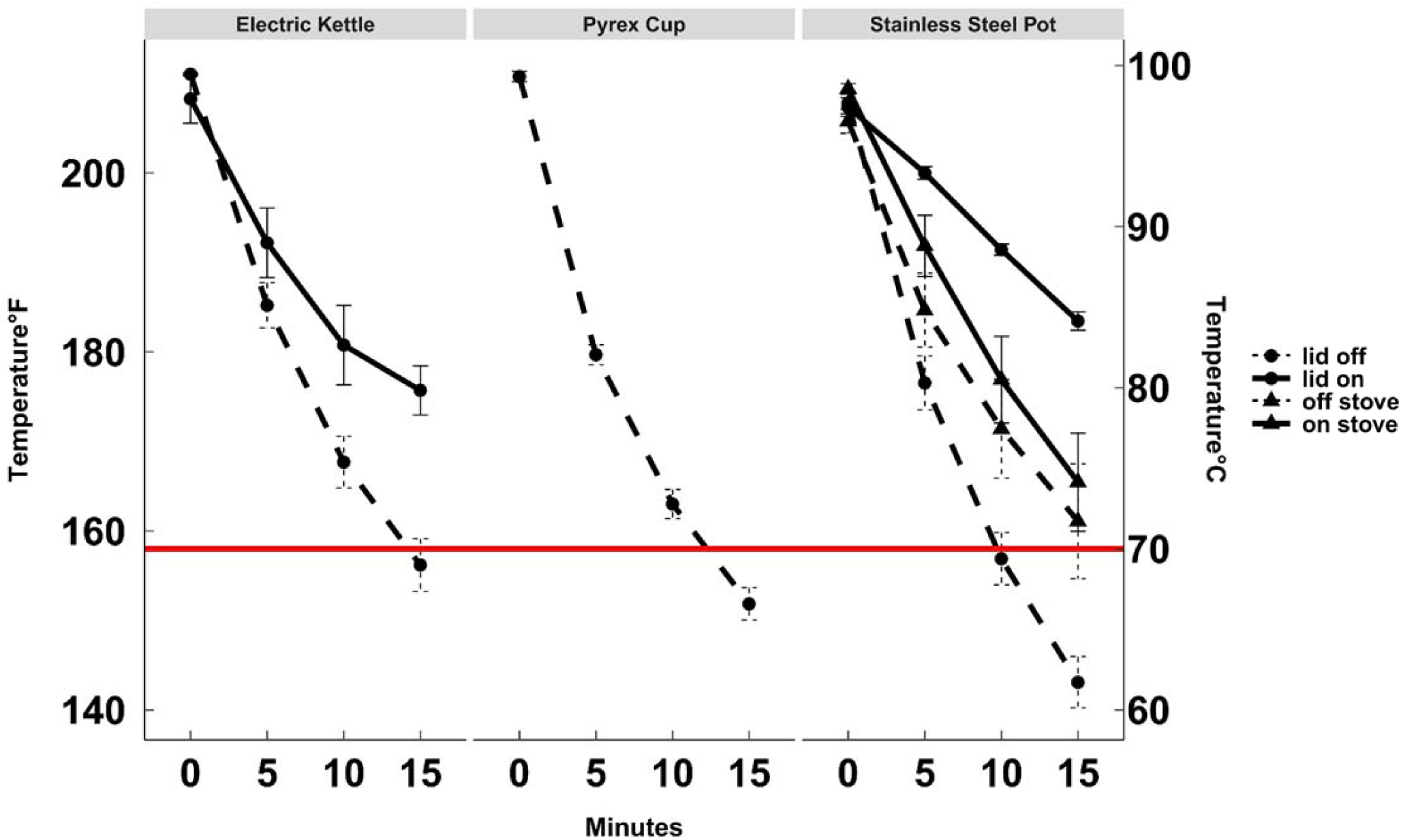
Water temperature (1,000 mL) in three vessel types (Electric Kettle, Pyrex Cup, Stainless Steel Pot) over 15 m. Circles indicate whether the vessel was covered with a lid. Triangles indicate whether the vessel was maintained on the heating surface. The “Off” (stove) condition is a dashed line and the “on” (stove) condition is a solid line, and it applies only to the stainless steel pot vessel.

When vessels did not have a lid, the water temperatures dropped below 158°F (70°C) between 5 and 15 min of cooling. Water temperatures in the Pyrex cup and stainless steel pot (lid off) dropped below 158°F (70°C) after 15 min of cooling in both volumes (Figure 1). Water temperatures in the electric kettle without a lid fell below 158°F (70°C) after 15 min of passive cooling. The lowest average water temperature of all preparation conditions 127.6 ± 1.79°F (53.1 ±0.98°C) occurred in the stainless steel pot at 15 min (625 mL water, lid off, off heating source). When lids were left on, water temperatures remained above 158°F (70°C) even after 15 min (Figure 2). The highest average water temperature at 15 min of all preparation conditions occurred in the stainless steel pot, 191 ± 1.82°F (88.3 ± 1.01°C) at 15 min (625 mL water, lid on, on heating source). Leaving the lid on could limit evaporation and interactions with the ambient environment (e.g., room temperature, relative humidity) that accelerate cooling. Notably, water temperatures ≥180°F (80°C) could compromise thermolabile nutrients (Agostoni, 2004). Leaving the vessel on the heating surface, after turning it off, also affected the rate of cooling. A warm stove can radiate heat that impacts water temperatures in the vessel (Datta, 2002). Therefore, the caregivers’ actions are important to consider when developing instructions for formula reconstitution.

Caregivers will likely use a variety of vessel types, water volumes, and heating sources for PIF preparation. Caregivers will also engage in a variety of practices for heating, such as removing or adding lids to the heating vessel or keeping or removing vessels on heating surfaces, or transferring the heated water to different containers. Each of these differences may impact water temperature. Federal guidance shows different vessels and preparation conditions within their infographics (CDC, 2024; CFR, 2016). For example, the CDC infographic shows a pot to boil water, but then shows a measuring cup, to transfer heated water into baby bottles. The Code of Federal Regulations (21 CFR § 107.20 subsection B) for PIF depicts a kettle to boil water on the stove, and then a measuring cup to transfer water to the baby bottle. Furthermore, regardless of the water temperature after 5 min of passive cooling, water temperature will continue to drop after transfer from the heating vessel to a baby bottle and addition of PIF.

Since caregivers may not strictly monitor cooling times (Malek, 2020), cooling periods could extend past the recommended “wait 5 minutes”. It is reasonable to assume that caregivers preparing PIF while caring for an infant may apply a holding period that extends longer than 5 min. Additionally, a common misconception is that the recommendation to heat water for PIF reconstitution exists to treat (sterilize) the water, not the PIF (Wilkinson, T, 2019; Malek, 2020). Hence, some caregivers boil, cool, then store water for future PIF reconstitution. These misinterpretations may be reinforced by guidelines and label instructions from imported formulas from Europe, in which PIF products reference “waiting ∼30 min” following boiling (NHS, 2023). Further reinforcement of this idea may come from FDA guidance that suggests modified water handling practices depending on the safety of the household’s water source (FDA, 2024), again emphasizing water safety. Although thermal treatment is a risk-reduction strategy for *Cronobacter* spp. in water too, PIF is not a sterile product and hot water reconstitution is recommended for high-risk infants to help mitigate risk that may be coming from PIF (Lui et al., 2013; Pina-Perez et al., 2016; Redmond & Griffith., 2013).

### Water temperature significantly decreased following transfer to the baby bottle

After 5 min of cooling in the heating vessel, water is then transferred to the baby bottle for PIF reconstitution. The water temperature prior to transfer, bottle material and capacity (i.e., size), and water volume used in reconstitution significantly impacted water temperatures once transferred to baby bottles (*p*<0.001). The highest average water temperature (188.2±2.5°F, 86.8±1.4°C) was recorded in 8 oz plastic bottles filled to capacity (i.e., with 8 fl. oz of water) at an initial temperature (i.e., in the heating vessel before transfer) of 203°F. The lowest average water temperature (149.3±0.9°F, 65.2±0.5°C) was recorded in glass, half-filled 4 oz bottles at an initial temperature of 165°F (Table 2). Plastic baby bottles always had higher water temperatures than glass bottles, regardless of initial water temperatures, bottle volume, and water volume (Table 2). Water temperatures in glass bottles were up to 30°F lower than in plastic bottles following transfer (Table 2). Glass has a higher thermal conductivity (1W/m*K) than plastic (∼0.3-0.4 W/m*K) (TheEngineeringToolbox, 2003), facilitating the heat loss from glass bottles to the surrounding environment. Increasing material thickness could also limit the heat transfer through the bottle wall (Kakaç et al., 2018).

Notably, water temperatures in half-filled bottles (e.g., 2 oz water in 4 oz bottle) were significantly lower than in full bottles (*p*<0.05), except in plastic bottles at 165°F initial water temperature. Water temperatures were up to 9°F lower in half-filled bottles than full bottles. The increased headspace in half-full bottles can facilitate ambient air flow into the bottle, which could increase heat loss from the water to the air from natural convection. Air has greater thermal diffusivity than water, so temperatures change more rapidly in air (Datta, 2002). Fill capacity also effects the surface area to volume ratio.

Nonetheless, initial water temperature at transfer was greatest predictor of whether water temperature met or exceeded the recommended temperature 158°F (70°C) in the baby bottle. Regardless of other conditions, the average water temperature in each baby bottle fell below 158°F when the initial water temperature was 165°F (73.8°C) and remained above 158°F when the initial temperature was 203°F (95.1°C). At an initial temperature of 165°F (73.8°C), the average water temperatures in the baby bottles dropped to between 149.3°F and 157.8°F (65.2°C and 69.9°C) (Table 2). At an initial temperature of 203°F (95.1°C), the average water temperatures in the baby bottles dropped to between 165.7°F and 188.2°F (74.2°C and 86.8°C) (Table 2). Within these initial temperature groups, there was variation based on bottle attributes.

Significant temperature decreases when smaller volumes of water are being transferred to baby bottles may be relevant for caregivers with newborns who generally require smaller volumes of formula for feeds. There are several scenarios where smaller volume bottles had water temperatures that fell below 158°F (70°C). The volume of water and headspace in the bottle will likely impact temperature change of the water following transfer. In the initial validation study conducted by Edelson-Mammel and Buchanan (2004) in which 158°F (70°C) was shown to provide inactivation of *Cronobacter* spp. (*E. sakazakii*) during PIF reconstitution in bottles, 180 mL (6 oz) of water was used to prepare PIF in a plastic bottle. These conditions likely minimized the temperature decreases that occur with smaller water volumes or when using glass bottles. This study also conducted a series of submerged coil experiments to generate *D-* values for 12 *Cronobacter* spp. (*E. sakazakii*) isolates. The most thermostable isolate had a *D_70°C_* = 3.9 s suggesting that a treatment with hot water at 158°F (70°C) is effective at *Cronobacter* spp. inactivation. However, ensuring the reconstitution performed by caregivers actually delivers water treatments at 158°F (70°C) is a challenge that must be addressed through effective instructions.

### The addition of PIF to the water in the baby bottle further decreased water temperature

After PIF addition, temperatures in the bottles dropped on average an additional 5.8°F (Table 3). Baby bottle capacity, water volume, initial water temperature, and bottle material significantly impacted temperatures at PIF addition (*p*<0.01) (Table 4). For the “best case” scenario (i.e., plastic full-capacity bottles) the temperature achieved after shaking was 179.5±1.6°F (81.9±0.9°C). For the “worst case” scenario (i.e., glass half-capacity bottles) the temperature achieved after shaking was 138.0±1.3°F (58.9 ± 0.7°C) (Table 3). Temperatures dropped a total of 23.5°F (13.2°C) and 27°F (14.9°C) from the initial high (203°F; 95.1°C) and low (165°F; 73.8°C) water temperatures in the heating vessels, respectively. This could be attributed to the thermal energy (heat) from the water transferring to the powder and dissolving it, which decreases the water’s temperature. The addition of a solute (PIF) to the water may also decrease the liquid’s heat capacity, increasing the cooling rate of the liquid (Datta, 2002). These results indicate that water in many preparation scenarios may not be hot enough to inactivate *Cronobacter* spp. in PIF.

**Table 4.**
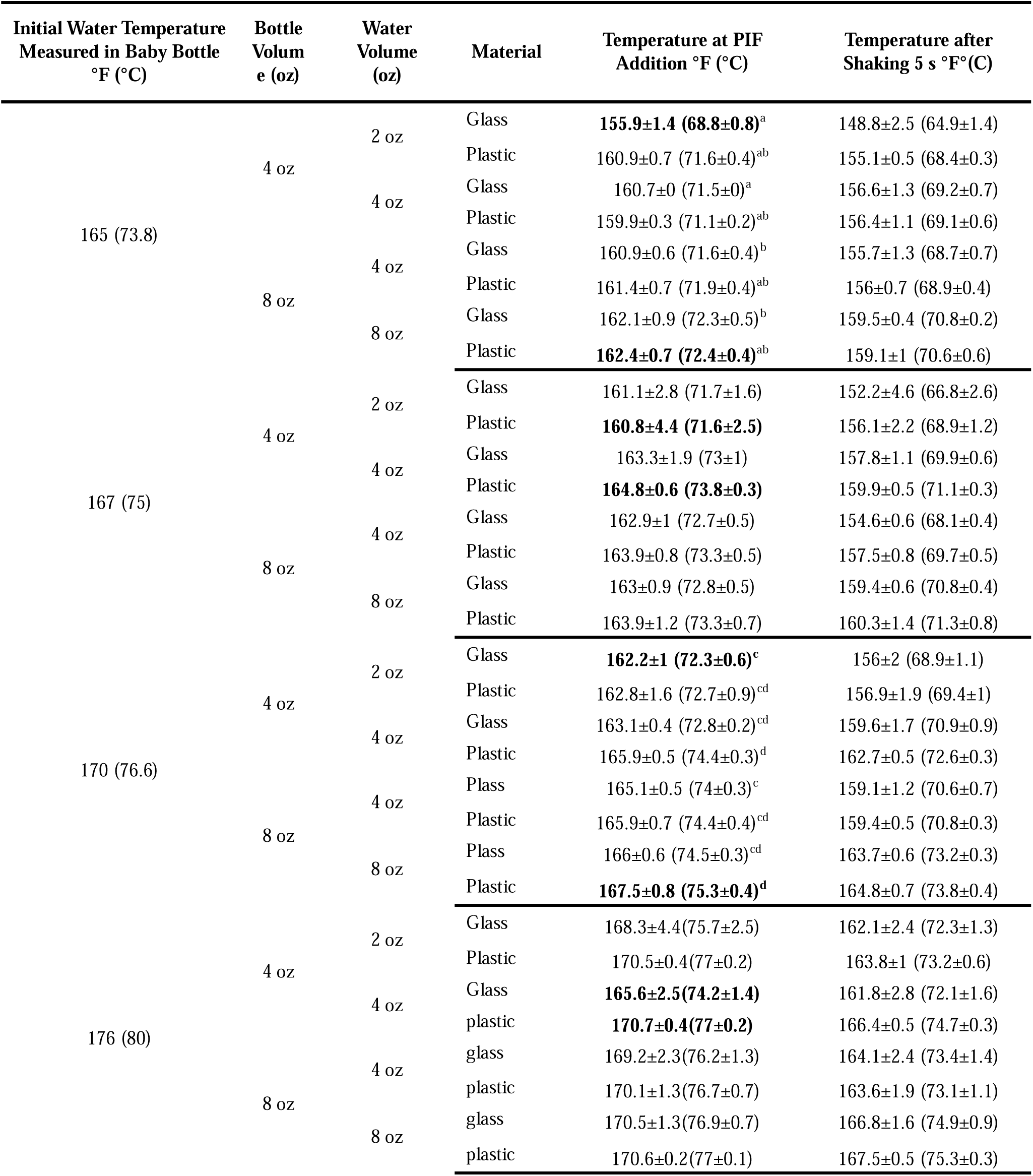
Temperatures during PIF reconstitution with hot water and shaking (Temperature Drop 3 and 4). Initial water temperatures in this experiment were pre-determined based on possible temperatures caregivers could monitor in the baby bottle before PIF addition. Results in bold text represent conditions that yielded the highest and lowest water temperature.

We asked the question, what initial water temperature measured in the baby bottle would ensure that prepared formula would meet or exceed the temperature of 158°F (70°C) following PIF addition. The temperature of prepared formula exceeded 158°F (70°C) when the initial water temperature in the baby bottle was recorded at 165°F (73.8°C), except for one condition (half-filled, 4 oz glass bottle, 165°F initial water temperature) (Table 4). These conditions resulted in formula at 155.9±1.4°F (68.8 ± 0.8°C), approximately 2.1°F below the recommended temperature. For formula prepared using 165°F water in baby bottles, temperatures following PIF introduction ranged from 155.9±1.4°F (68.8 ± 0.8°C) to 162.4±0.7°F (72.4±0.4°C). Increasing the initial water temperatures at 167°F resulted in temperatures approximately 5°F (2°C) warmer than those of 165°F (70°C) after PIF addition. The additional benefits to microbial safety should be balanced against risk of nutrient degradation and burns at increasingly elevated (Agostoni, 2004).

Water temperature can impact how PIF dissolves in water. Notably, water at a temperature greater than 140°F (60°C) may rapidly hydrate the outer powder particles of PIF, resulting in gel-like clumps that prevent complete dissolution (Figure 3). This may slow the dissolution process, limit mixing, and increase clumping (Walstra et al., 2005). Additionally, this incomplete homogenization may protect pathogens on the inside of these clumps from thermal inactivation. Mechanically mixing the water and powder for 5 s increases the wettability of the powder and improve its solubility (Fitzpatrick, 2016). Therefore, water temperature during PIF addition should not far exceed 158°F (70°C) to reduce physical changes that reduce ease of mixing. Moreover, even temperatures below 158°F (70°C) may still reduce *Cronobacter* spp. appreciably even if they do not achieve a 5 log reduction. This principle should be further explored through risk assessments to evaluate the tradeoffs between nutrient degradation and clumping with pathogen inactivation, given the likely low concentration of *Cronobacter* spp. typically present in contaminated PIF (Jongenburger et al., 2011).

**Figure 3.**
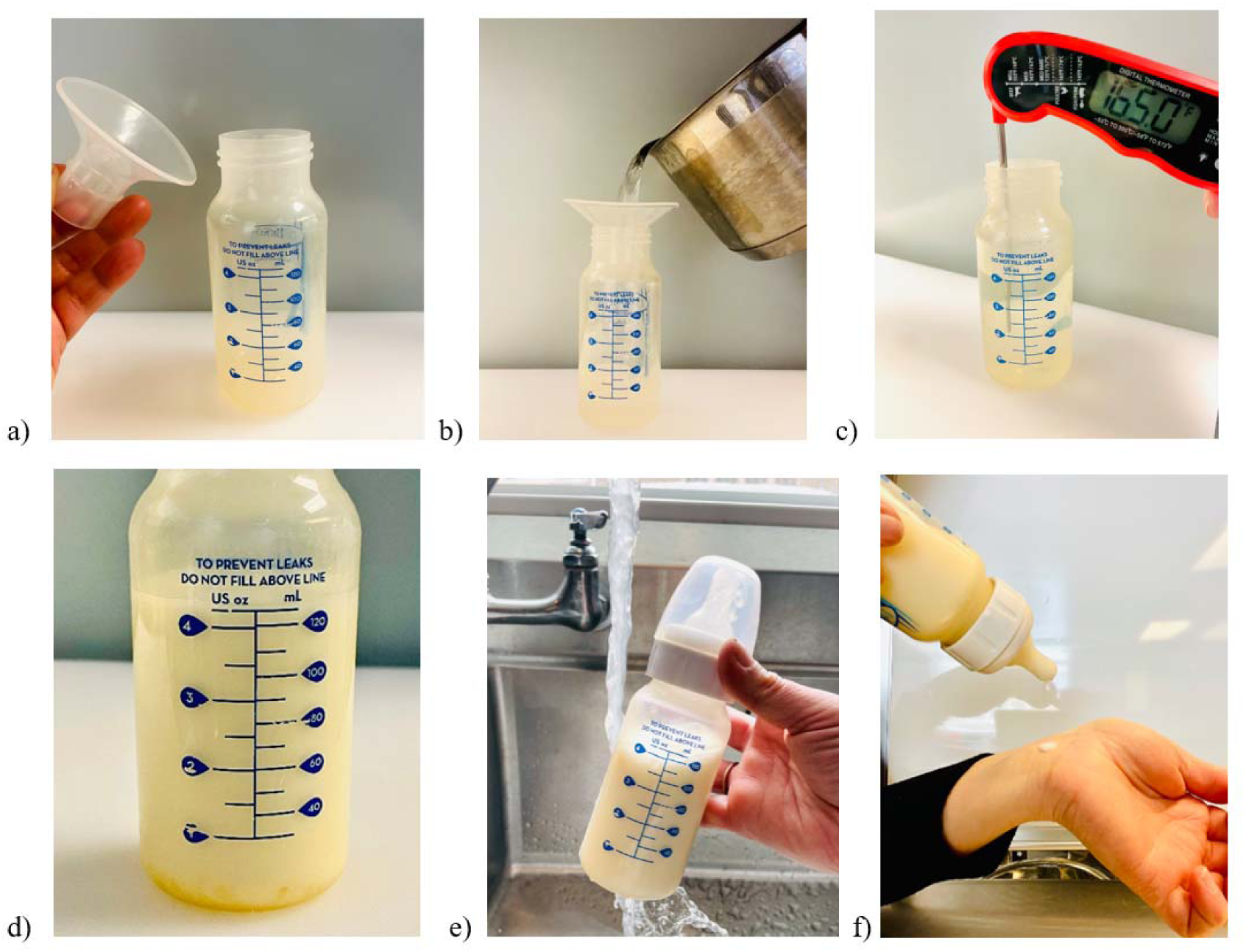
Steps in PIF reconstitution using hot water. Pictures show the sequence of actions (a) using a breast pump flange as a funnel (b) while pouring hot water into the baby bottle and (c) checking the temperature prior to PIF addition. Immediately after PIF addition to hot water, (d) PIF clump form on the bottom of the bottle before being broken up during the shaking step. After shaking, (e) the baby bottle is held under running water until (f) it is cooled to body temperature as confirmed by checking droplets on the wrist.

The hotshot method resulted in formula temperatures that were well below 158°F (70°C). Temperatures ranged from 120.6±5.3°F (49.2±2.9°C) to 140.3±1.1°F (60.2±0.6°C) following PIF addition (Table 3). Temperatures then decreased to between 93°F (33.8°C) and 113°F (45°C) after the addition of room temperature tap water (Table 3). The addition of only 1 fl. oz of 158°F (70°C) water is unlikely to sufficiently thermally treat PIF during reconstitution. Even though this method is quick, and most likely adapted from automated formula making machines (e.g., Baby Brezza), this reconstitution approach may have limited thermal control and more likely serves to warm formula to body temperature for consumption.

Preparation instructions that include transferring hot water increases the risk of burns to the caregiver preparing the bottle. However, it is likely similar to the risk associated with other recommendations around sterilizing bottle parts via a boiling water bath (CDC, 2024) or opening steaming commercial sterilizing bags from the microwave. A wide funnel used exclusively for formula preparation, or even a clean breastfeeding flange, would be practical aids for pouring hot water into the baby bottle (Figure 3). Additionally, caregivers may perceive temperature monitoring as burdensome. However, willingness to perform these tasks could increase if caregivers are aware of the risk of *Cronobacter* spp. infection among high-risk infants and provided with a clear temperature recommendation. It is also important to note that hot water preparation instructions generally apply to infants only in their first 2 months of life, reducing the overall burden of this task on caregivers. Existing educational strategies regarding temperature monitoring in food preparation could be leveraged to communicate with caregivers about PIF preparation.

### Cooling times of reconstituted PIF vary depending on the method

Methods for cooling prepared formula to body temperature (∼37°C) varied in duration. CDC guidance recommends caregivers “wait for the formula to cool” until it is “warm, not hot,” citing body temperature as an appropriate indicator (CDC, 2024). Running water over a glass bottle (half-filled, 4 oz bottle size) resulted in the shortest overall cooling time required to reach body temperature at ∼1.4 min, which was approximately 1.9 and 23.1 min faster than cooling with an ice bath or with passive cooling at room temperature, respectively. Running water over a plastic bottle (filled with 8 oz formula, 8 oz bottle size) cooled reconstituted formula to body temperature in 6.8 min, which was 4.9 and 51.5 min faster than the ice bath or passive cooling at room temperature, respectively. Leaving the bottle at room temperature resulted in the longest overall cooling duration at 58.3 min (full 8 oz plastic bottle) and 24.5 min (half-filled 4 oz glass bottle). Short cooling durations were observed in glass bottles filled to half capacity, which coincides with the prior thermal trends achieved after PIF addition and shaking (Figure 4). The peaks observed on the cooling curve of a full plastic 8 oz bottle via ice bath resulted from the process of “swirling” performed to increase blending of the colder portion of the liquid (submerged in the ice bath), with the warm portion (at room temperature, given the height of the bottle) (Figure 3 and 4).

**Figure 4.**
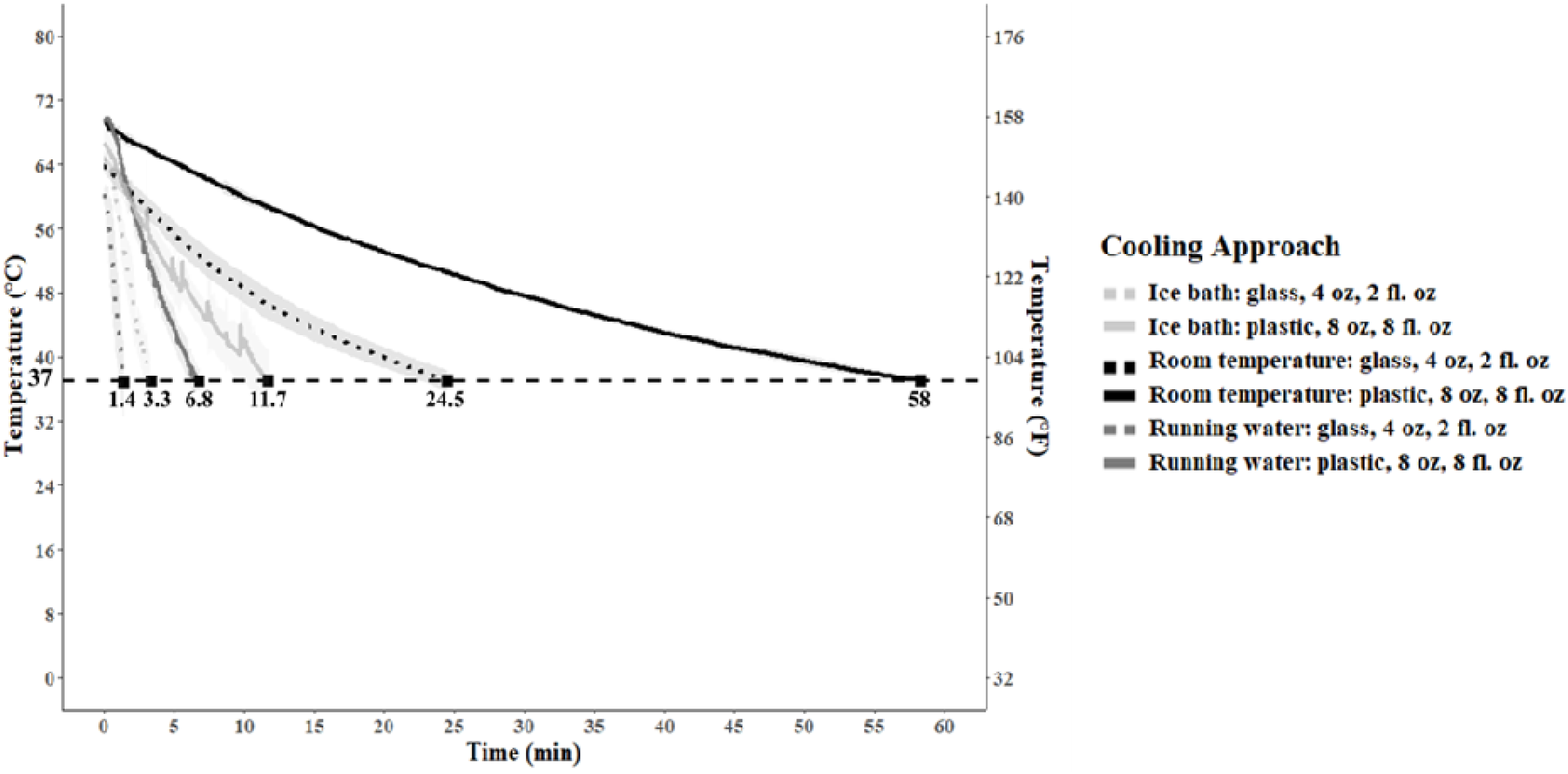
Temperature of reconstituted formula in baby bottles during the period of cooling to approximately body temperature. Cooling curves are the average of three replicate experiment for two types of bottles representing the fastest and slowest cooling dynamics: (1) 4 fl. oz glass bottles filled with 2 fl. oz of formula and (2) 8 fl. oz plastic bottles filled with 8 fl. oz of formula. Three cooling treatments were evaluated for each type of bottle: (1) passive cooling at room temperature, (2) holding the bottles under cold running water, and (3) submerging bottles in an ice bath. Note that the spikes in the curve for the ice bath treatments for 8 fl. oz bottle correspond to points where the bottle was swirled to mix contents as the ice bath only reached about half the height of the bottle.

Plastic bottles and minimal headspace may keep water temperatures high for longer durations, which could further enhance C*ronobacter* spp. inactivation during cooling. However, below temperatures that are lethal, holding formula for too long in the “danger zone” temperature range (40 to 140°F, 4 to 60°C) may eventually lead to *Cronobacter* spp. recovery. For example, in a work of Chen et al (2009) reconstituted infant formula held at 50°C supported *Cronobacter* spp. survival for at least 3 h (Chen et al, 2009). The WHO (2012) states that reconstituted PIF should be stored at temperatures ≤ 5°C to prevent the growth of *Salmonella* and *C. sakazakii* (previously, *E. sakazakii*). Furthermore, prolonged cooling times are impractical for caregivers as delayed feedings may induce emotional distress for the infant. In practice, even a cooling duration of approximately 6.8 min (running water, full 8 oz plastic bottle) is a long time for a caregiver to actively hold the bottle under a faucet. Caregivers, especially if they are inexperienced in formula preparation, may shorten the cooling step to appease a hungry infant. However, high temperatures could scald or burn infants, compromising their health, and inducing feeding aversion. Notably, Remond and Griffith (2013) reported that 60% of observed caregivers (n=50) did not check temperatures of reconstituted formula prior to feedings. Although the CDC (2024) recommends testing the temperature of the prepared formula “by putting a few drops on your wrist,” sensitivity to pain (i.e., heat) varies among caregivers. Chiang et al. (2023) reported that 1 to 4 PIF-related burn cases occurred per year (between 2017-2019) in U.S. hospitals, a likely underestimate of the true burden of burn injuries related to hot formula. Redmond and Grifith (2013) reported that recruited caregivers cooled bottles via running water (20%) and cold water in a basin (60%) in 5 to 11 min and 1 to 3.5 min, respectively, which is similar to the findings reported here. Caregivers are also cautioned to avoid water introduction into the nipple or bottle during active cooling to prevent recontamination of the prepared formula (CDC, 2024).

### The labels on PIF containers evaluated in this study did not describe temperature and time recommendations for caregivers of high-risk infants

Only three products from one European brand recommended a water temperature of 70°C (158°F) to prepare formula. Some brands (20/36, 56%) suggested using cooled, boiled water, while others (2/36, 6%) recommended warm previously boiled water, for reconstituting PIF. A temperature recommendation of 40°C (100°F) was found in 11 products, presumably related to protecting the probiotic cultures contained in the PIF. Formula manufacturers are not required to add probiotic cultures to their PIF products, nor can they promote formula benefits in a way that challenges breastfeeding per the International Code of Marketing of Breast-Milk Substitutes (UNICEF, 2023). Furthermore, in a report made by the European Food Safety Authority (EFSA, 2014) the authors reviewed a number of studies and systematic reviews on the potential benefits of probiotics in infant formula, and did not find any significant physiological or health effects among those consuming supplemented formula compared with those given unsupplemented formula (Crawley et al, 2022). Inconsistencies surrounding water temperature and heating recommendations may confuse caregivers. All PIF products mentioned healthcare providers, stating “Ask your baby’s doctor (or healthcare provider) about the need to use cooled boiled water for mixing”, and the stated caregivers should “boil (sterilize) bottles, nipples, and rings before use.” The CFR 21 § 107.20 subchapter B includes requirements for PIF labels, including statements about sterilization of water, bottles, and nipples (CFR, 2024). Although this regulation shows an infographic where hot water is used in reconstitution, the prescribed language defers to physicians for the final decisions about sterilization of equipment rather than providing instructions on the label at point of use.

All PIF product labels contained the same recommendations regarding storage and temperature conditions of prepared, unfed formula. These recommendations included storing prepared, unfed formula ≤24 h at 2°C to 4°C, and that an open can of formula should not be kept for more than a month (Table 5). Less than half of labels (15/36, 42%) clearly mentioned that prepared, unfed formula should remain unrefrigerated ≤ 2 h. Most brands (35/36, 97%) indicated a one hour limit for leftover formula after initiating feeding, and one indicated discarding the feed right away. Only one brand mentioned the safety of the water source. All labels provided a warning to not microwave formula. Accidental injuries can occur when overheating infant formula in a microwave oven, including scald burns of the trachea, palate, and oropharynx due to aspiration and ingestion of foods that have been overheated (Dixon J., 1997). One brand (three products) indicated caregivers should check formula temperatures on the inside of their wrist (i.e., dropping a few droplets of ready to feed formula) before feeding the baby (NHS, 2023; FDA, 2024) (Figure 3).

**Table 5.**
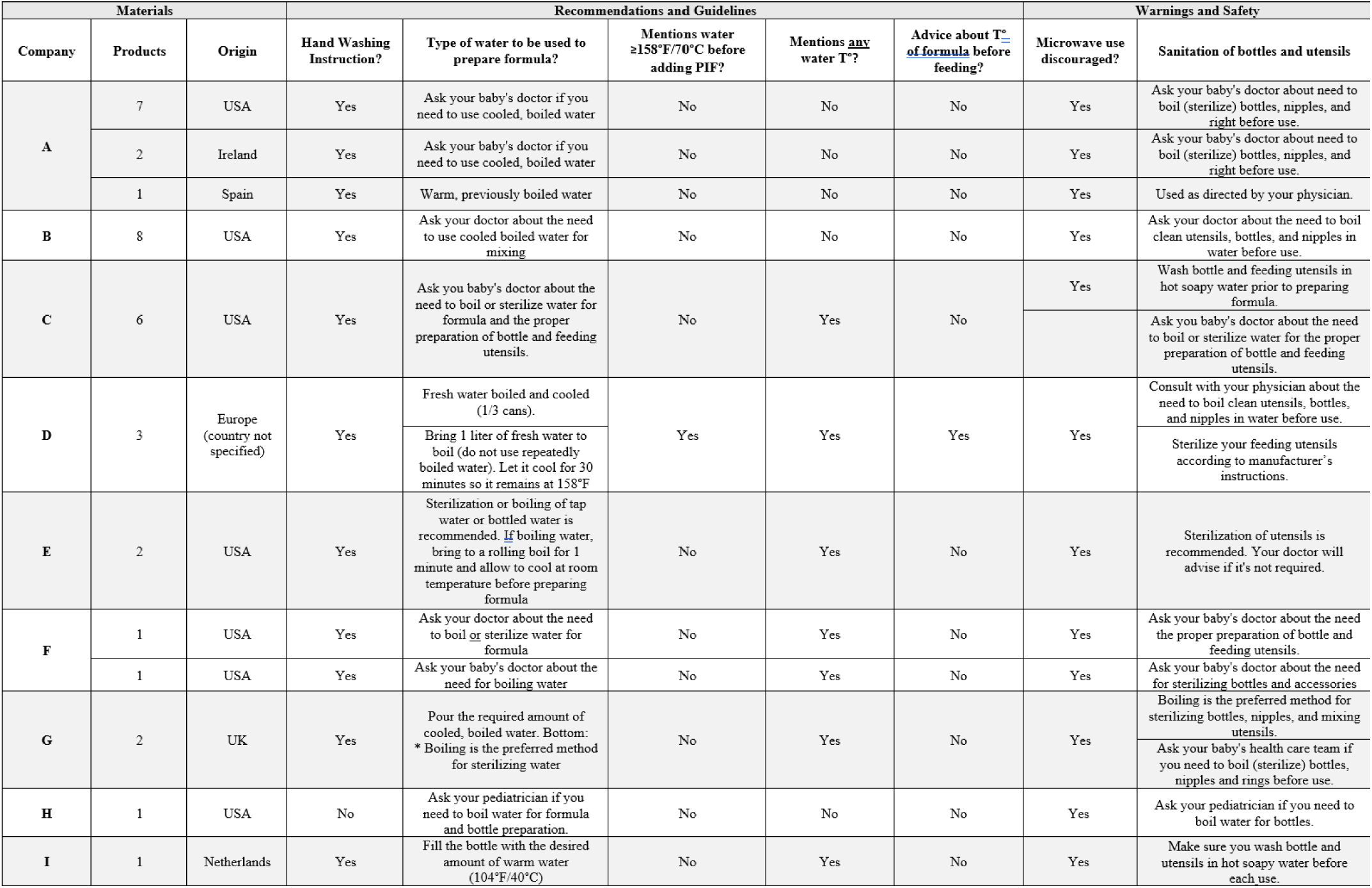
Assessment of safe handling instructions on PIF labels (n=36) available in the U.S. market from 9 different brands. T°= temperature.

| Materials |  |  | Recommendations and Guidelines |  |  |  |  | Warnings and Safety |  |
| --- | --- | --- | --- | --- | --- | --- | --- | --- | --- |
| Company | Products | Origin | Hand Washing Instruction? | Type of water to be used to prepare formula? | Mentions water $\geq 158^{\circ}\text{F}/70^{\circ}\text{C}$ before adding PIF? | Mentions any water T°? | Advice about T° of formula before feeding? | Microwave use discouraged? | Sanitation of bottles and utensils |
| A | 7 | USA | Yes | Ask your baby's doctor if you need to use cooled, boiled water | No | No | No | Yes | Ask your baby's doctor about need to boil (sterilize) bottles, nipples, and right before use. |
|  | 2 | Ireland | Yes | Ask your baby's doctor if you need to use cooled, boiled water | No | No | No | Yes | Ask your baby's doctor about need to boil (sterilize) bottles, nipples, and right before use. |
|  | 1 | Spain | Yes | Warm, previously boiled water | No | No | No | Yes | Used as directed by your physician. |
| B | 8 | USA | Yes | Ask your doctor about the need to use cooled boiled water for mixing | No | No | No | Yes | Ask your doctor about the need to boil clean utensils, bottles, and nipples in water before use. |
| C | 6 | USA | Yes | Ask you baby's doctor about the need to boil or sterilize water for formula and the proper preparation of bottle and feeding utensils. | No | Yes | No | Yes | Wash bottle and feeding utensils in hot soapy water prior to preparing formula. |
|  |  |  |  |  |  |  |  |  | Ask you baby's doctor about the need to boil or sterilize water for the proper preparation of bottle and feeding utensils. |
| D | 3 | Europe (country not specified) | Yes | Fresh water boiled and cooled (1/3 cans). | Yes | Yes | Yes | Yes | Consult with your physician about the need to boil clean utensils, bottles, and nipples in water before use. |
|  |  |  |  | Bring 1 liter of fresh water to boil (do not use repeatedly boiled water). Let it cool for 30 minutes so it remains at 158°F |  |  |  |  | Sterilize your feeding utensils according to manufacturer's instructions. |
| E | 2 | USA | Yes | Sterilization or boiling of tap water or bottled water is recommended. If boiling water, bring to a rolling boil for 1 minute and allow to cool at room temperature before preparing formula | No | Yes | No | Yes | Sterilization of utensils is recommended. Your doctor will advise if it's not required. |
| F | 1 | USA | Yes | Ask your doctor about the need to boil or sterilize water for formula | No | Yes | No | Yes | Ask your baby's doctor about the need the proper preparation of bottle and feeding utensils. |
|  | 1 | USA | Yes | Ask your baby's doctor about the need for boiling water | No | Yes | No | Yes | Ask your baby's doctor about the need for sterilizing bottles and accessories |
| G | 2 | UK | Yes | Pour the required amount of cooled, boiled water. Bottom: * Boiling is the preferred method for sterilizing water | No | Yes | No | Yes | Boiling is the preferred method for sterilizing bottles, nipples, and mixing utensils. |
|  |  |  |  |  |  |  |  |  | Ask your baby's health care team if you need to boil (sterilize) bottles, nipples and rings before use. |
| H | 1 | USA | No | Ask your pediatrician if you need to boil water for formula and bottle preparation. | No | No | No | Yes | Ask your pediatrician if you need to boil water for bottles. |
| I | 1 | Netherlands | Yes | Fill the bottle with the desired amount of warm water (104°F/40°C) | No | Yes | No | Yes | Make sure you wash bottle and utensils in hot soapy water before each use. |

### Other safe handling recommendations in the preparation of infant formula

Several other safe-handling recommendations on PIF labels were recorded. Advice for handwashing was noted (e.g., written text, drawings, identifying soap/water, etc.). From the 36 PIF labels analyzed, 97% (35/36) recommended hand washing. Soap and water were identified in 44% (16/36) of the labels, but water temperature recommendations were limited (some just mentioning using warm water) or omitted entirely. A drawing or picture of hand washing did not always accompany the recommendations on the labels (19/36). The word “thoroughly” (to emphasize the action to be done exhaustively and completely) was used in 28% (10/36) of hand washing recommendations.

Handwashing is important for maintaining the microbial safety of prepared formula. This step could reduce the risk of formula cross-contamination via caregivers’ hands. This is especially relevant if the formula scoop is submerged in PIF, requiring the caregiver to touch the product to extract the scoop. We noticed that the scoops included in PIF cans were not always accessible (e.g., adhered to the lid), prompting caregivers to “dig” into the infant powder. Bacteria could persist in PIF if introduced from hands, a spoon, etc.

Font size within label instructions was quite small. Letters were approximately 1 to 2 mm, which is the minimum size recommended for the display of information in labels (Food Standards Agency, 2023). Furthermore, the International Code of Marketing for Breast-Milk Substitutes specifically recommends clarity in preparation instructions (WHO, 1981). While some PIF labels provided contact information for communication with “feeding experts” (available during regular business hours), these additional steps, similar to consultation with a healthcare provider, place the burden on the caregiver to seek out clarification on when and how hot water reconstitution should be performed. In other instances, a QR code was provided on the cans, which routed to the company’s website. However, the website did not link directly to preparation instructions, but provided options to buy the product or check individual testimonies. Label attributes are a key resource for caregivers, and further consideration of their readability, content, and structure is warranted.

### Final considerations and opportunities for future work

Our study demonstrates how reasonably anticipated variation in formula preparation methods translates into differences in water temperature during PIF reconstitution. Based on the outcomes from the scenarios we tested, if caregivers strictly follow the current guidelines of “boil water and wait no longer than 5 min,” water temperatures may be well above 158°F (70°C), potentially compromising the nutritional profile of PIF, or well below this target temperature, decreasing *Cronobacter* spp. lethality. A target water temperature of 165°F (73.8°C) measured in the bottle prior to PIF addition generally provided a PIF treatment temperature at or above 158°F (70°C) during reconstitution. In this scenario, caregivers would use a probe thermometer to check water temperature in the baby bottle, then add PIF to the bottle. Similarly, future studies should investigate how these temperatures could impact thermolabile nutrients and PIF composition. Currently, there is not a consensus on how hot water reconstitution instructions should be presented to caregivers, or even whether they should be presented at all. However, ambiguous instructions that likely result in PIF reconstitution at insufficient temperatures to reduce *Cronobacter* spp. or directs caregivers to other sources of information poorly serve caregivers of high-risk infants.

## Data Availability

All data produced in the present study are available upon reasonable request to the authors

## Acknowledgements

This work was supported in part by the U.S. Department of Agriculture, National Institute of Food and Agriculture [Hatch Project 2024-25-191] to ABS.

## FIGURES AND TABLES

**Supplementary Figure 1.**
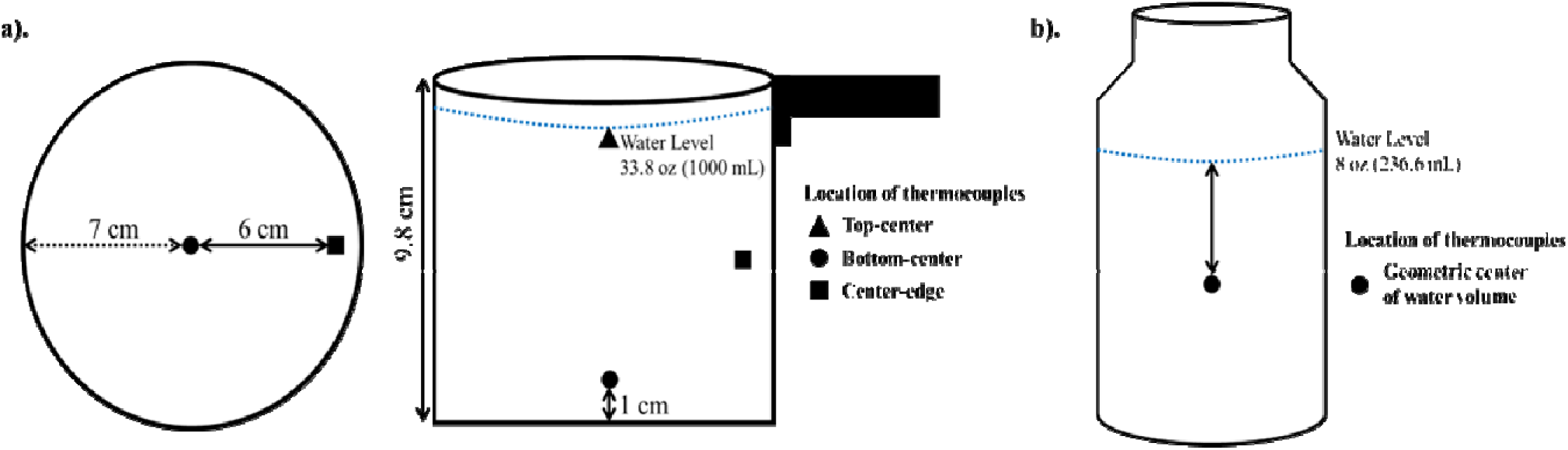
Representative schematic of locations of the K-type thermocouples in a). heating vessels (Left-Right: Diameter and height of stainless-steel pot) and b). baby bottles.

**Supplementary Table 1.**
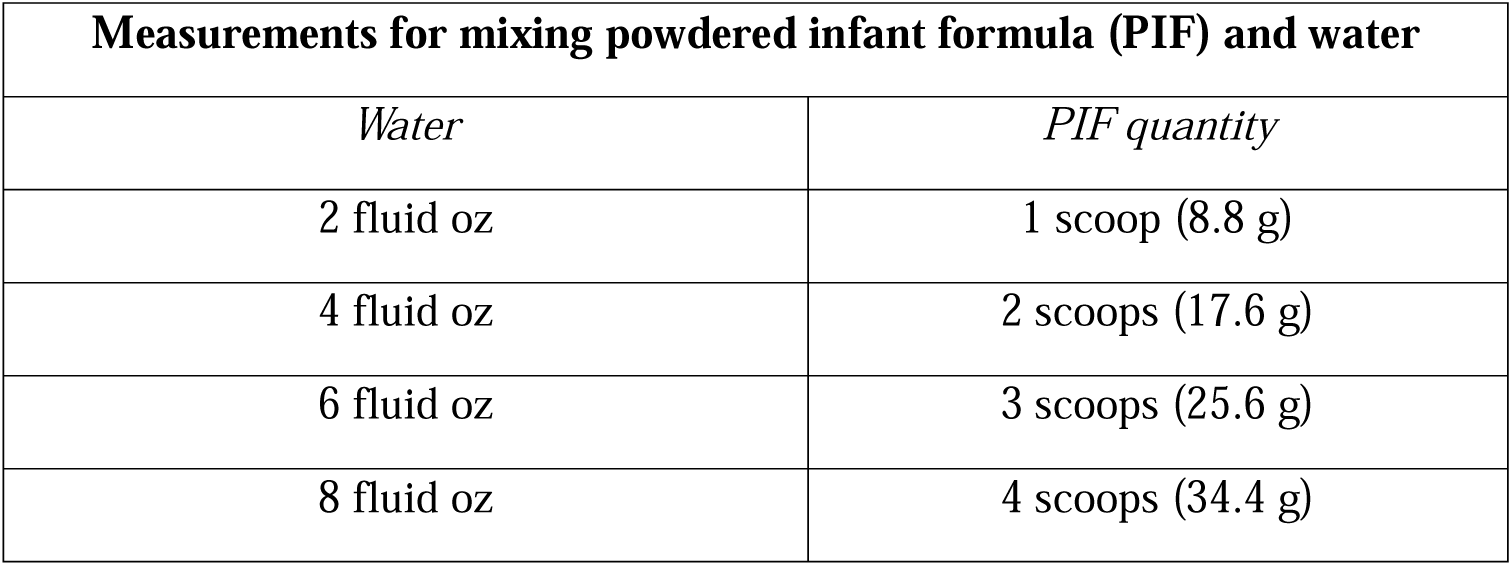
Standard reconstitution quantities of PIF per fluid ounces of water to prepare baby bottles.

